# Sample Size Calculations for Variant Surveillance in the Presence of Biological and Systematic Biases

**DOI:** 10.1101/2021.12.30.21268453

**Authors:** Shirlee Wohl, Elizabeth C. Lee, Bethany L. DiPrete, Justin Lessler

## Abstract

As demonstrated during the SARS-CoV-2 pandemic, detecting and tracking the emergence and spread of pathogen variants is an important component of monitoring infectious disease outbreaks. Pathogen genome sequencing has emerged as the primary tool for variant characterization, so it is important to consider the number of sequences needed when designing surveillance programs or studies, both to ensure accurate conclusions and to optimize use of limited resources. However, current approaches to calculating sample size for variant monitoring often do not account for the biological and logistical processes that can bias which infections are detected and which samples are ultimately selected for sequencing. In this manuscript, we introduce a framework that models the full process— including potential sources of bias—from infection detection to variant characterization, and we demonstrate how to use this framework to calculate appropriate sample sizes for sequencing-based surveillance studies. We consider both cross-sectional and continuous sampling, and we have implemented our method in a publicly available tool that allows users to estimate necessary sample sizes given a specific aim (e.g., variant detection or measuring variant prevalence) and sampling method. Our framework is designed to be easy to use, while also flexible enough to be adapted to other pathogens and surveillance scenarios.

## INTRODUCTION

The emergence of SARS-CoV-2 variants with different epidemiologic properties has contributed to difficulties in controlling the COVID-19 pandemic. Towards the end of 2020, the first Variant of Concern (VOC) ^1^, later designated Alpha, was identified in the United Kingdom ^2^, and additional VOCs have continued to be identified throughout the pandemic ^3–5^. By definition, VOCs are associated with increased SARS-CoV-2 transmissibility, increased virulence, or decreased effectiveness of available diagnostics, vaccines, or therapeutics ^6^. As VOCs may trigger large resurgent waves of disease or other substantial changes in pathogen epidemiology, early detection and tracking of variants is a critical component of pandemic response.

Whole genome sequencing of SARS-CoV-2 samples allows for detection of novel variants and regular monitoring of their frequency in populations. Although genomic sequencing has become faster and more cost-efficient, it is not possible or necessary to sequence clinical samples from all cases, and efficient allocation of resources (e.g., time and supplies) is critical for public health response. Therefore, selecting an appropriate subsample for sequencing should play an important role in VOC monitoring. However, sample sizes are still often dictated by cost and convenience, and there is limited guidance available for designing population sampling strategies for genomic studies ^7^, including surveillance efforts aimed at detecting and characterizing VOCs.

Initial attempts at sample size calculations for tracking of SARS-CoV-2 VOCs have focused on determining the number of samples needed to identify a variant at a particular frequency in the population ^8–11^. These frameworks often start at the point where SARS-CoV-2 samples are returned positive or start with assumptions about the proportion of SARS-CoV-2 infected individuals that are tested and detected. However, the composition of this pool of detected infections may be biased by variant-specific differences in transmissibility, case detection, and test sensitivity. Hence, there is a need for a more comprehensive framework that models the full process from infections to detected infections to sequenced samples. Furthermore, sampling strategies should consider the fact that variant surveillance is an ongoing process and that variant frequencies change over time.

Here we aim to develop an easy-to-use, actionable framework for selecting the appropriate number of samples for sequencing when the goal is (1) detecting new variants as they arise in a particular population or geographic region, or (2) measuring the prevalence of specific variants in that population. Although there are a number of limitations inherent to the approach described below, it is applicable to a wide variety of settings and requires estimation of only a limited number of parameters in order to obtain a reasonable estimate of the required sample size for monitoring pathogen variants. While our work focuses on genomic sequencing for SARS-CoV-2 variant identification, this general framework can be applied to other pathogens and molecular methods.

### Conceptual Framework and Approach

When designing a sequencing-based study or surveillance system, the first step is to identify the population of interest. The framework presented here can be applied to populations at any scale—such as a country, state, or community—as long as the required parameters (explained in detail in the next section) can be estimated for that population. Once the relevant population is identified, the next step is to determine the specific question(s) to answer, as different goals require different sample sizes in order to obtain reliable results. Our framework provides guidance for sample size calculation and bias correction for both variant detection and measuring prevalence ^12^ in the context of a single cross-sectional snapshot or an ongoing surveillance program (**Fig 1**). Sampling frequency will influence how sample size is defined (i.e., overall study size versus average daily or weekly sampling rate) and what targets must be specified to calculate the appropriate sample size. For instance, in a cross-sectional study, a possible target could be the probability of detecting a variant at a particular prevalence, while with ongoing surveillance it might be the waiting time to detect a recently introduced variant that is growing in prevalence. If the study design is fixed (i.e., a set number of samples have already been collected or sequenced), the same principles can be applied to evaluate our confidence in the results.

**Figure 1.**
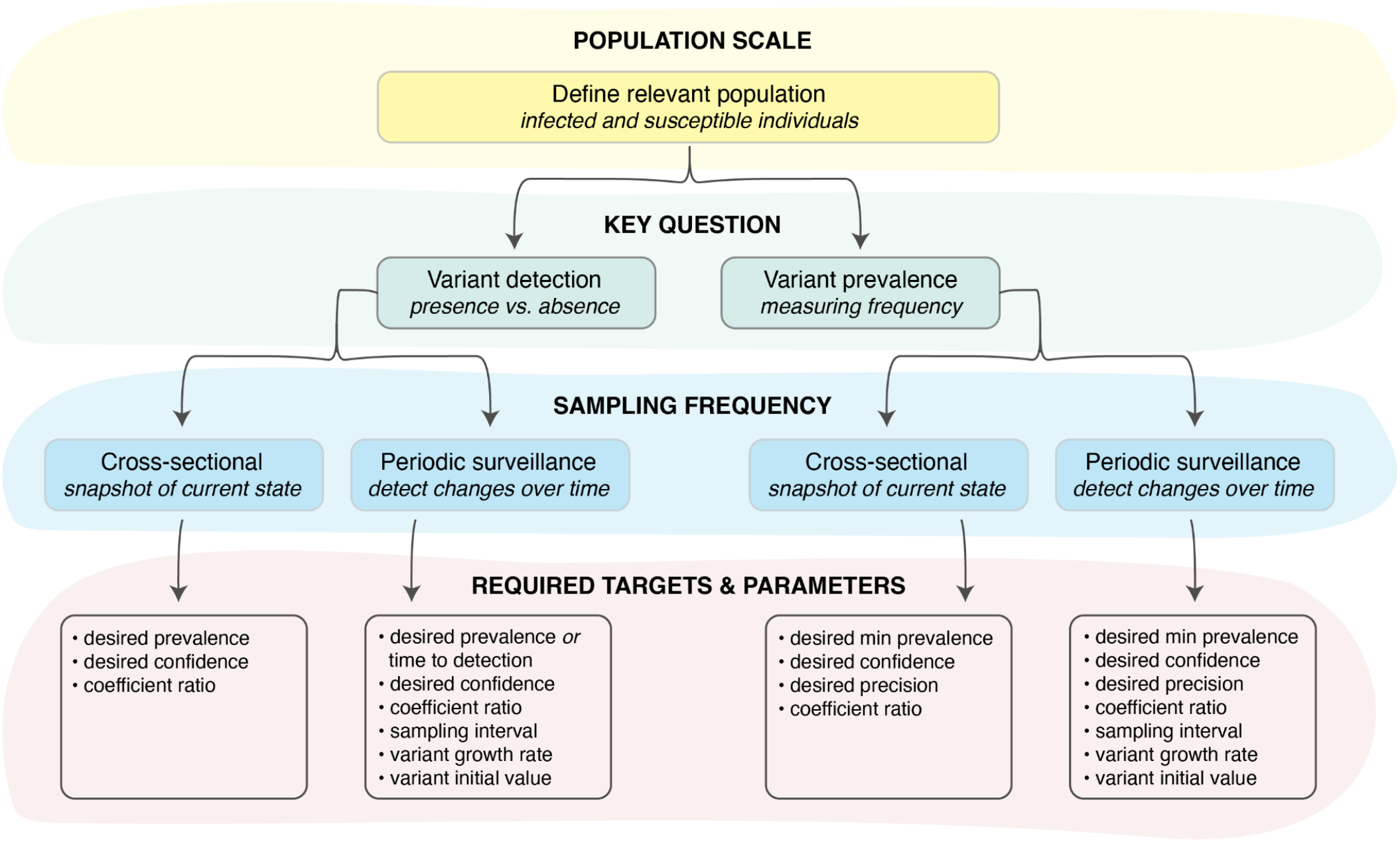
Decision tree for designing a variant surveillance program. Identifying the relevant population (yellow shaded region), key goals of the study (green shaded region; variant detection or measuring variant prevalence) and sampling method (blue shaded region; cross-sectional or periodic) are necessary to determine the required targets and parameters (red shaded region) that must be specified in order to calculate the appropriate sample size. Importantly, parameter estimates should be specific to the pre-defined population of interest as well as reflective of the logistical and biological sources of bias at the time of sampling.

### Limitations of existing sampling strategies

We can use existing sampling theory as the basis of our approach to variant detection and prevalence estimation. Specifically, the sample size needed to detect novel variants at some probability can be calculated with a simple application of the binomial distribution. The probability of detecting at least one case belonging to a specific variant (*V*_*i*_) given the prevalence of this variant in the population 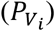 is equivalent to one minus the probability of not detecting it at all. Therefore, the sample size (*n*) needed to detect at least one case of a VOC at a pre-determined probability (*p*) has been shown to be ^10^:

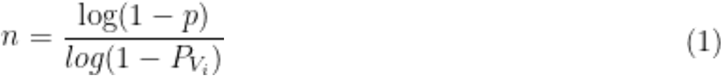

Similarly, existing sampling theory can be used to estimate the prevalence of known VOCs in a population. Specifically, sample size calculations for estimating proportions can be used to determine the number of sequences that should be generated to estimate VOC prevalence within a desired confidence interval ^13^:

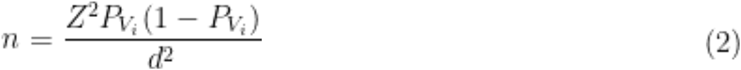

Where *n* is the number of samples needed, Z is the Z-statistic for the 95% confidence level, 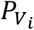 is the expected prevalence of the VOC in the population, and *d* is the desired absolute precision (tolerance for error in the prevalence estimate). This methodology has previously been used to calculate the number of SARS-CoV-2 samples needed to detect variants at different frequency levels ^8^ and it assumes that the sample size (*n*) is small compared to the total infected population.

These approaches to sample size calculation are subject to limitations. For one, both equations assume that the pool of samples available for sequencing is a representative random sample of the total infected population. However, the biology and epidemiology of SARS-CoV-2 VOCs, such as heterogeneity of disease severity, may affect which samples are collected and sequenced (**Fig 2, Fig S1**). The sequences used for analysis may therefore not be directly reflective of the underlying distribution of viral sequences, and this bias may be detrimental or useful depending on the goals of surveillance.

**Figure 2.**
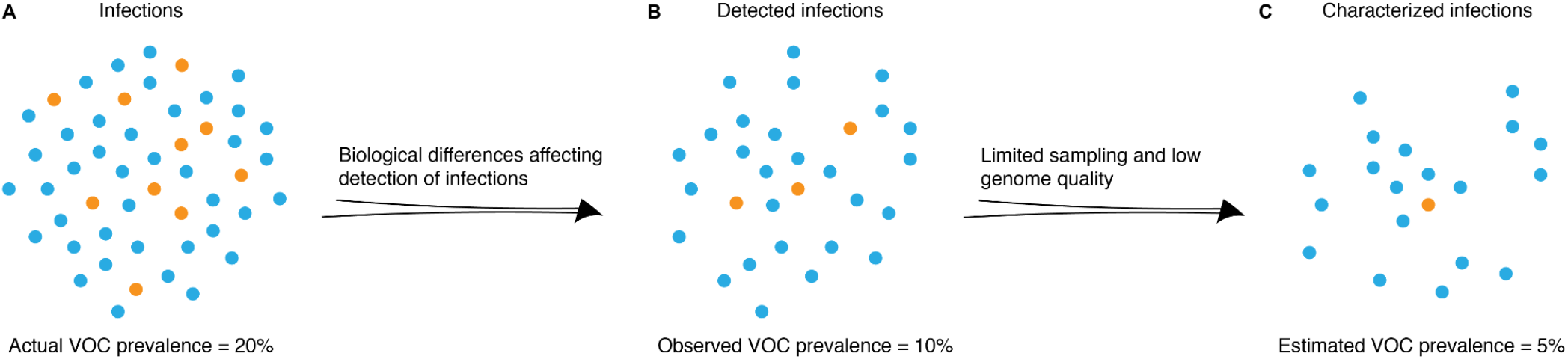
Factors affecting observed variant prevalence. VOC prevalence in (**A**) total population, (**B**) pool of detected infections, and (**C**) characterized infections (identified as a particular variant by sequencing or another technology). Biological and logistical differences between variants can lead to bias in observed variant proportions. Orange = infections caused by VOC (variant of concern); blue = infections caused by other variants of the same pathogen.

Here, we characterize the mechanistic process from infection to case detection to variant identification using a simple modeling framework that captures how these processes differ between variants. We first explore how VOC attributes could bias detection of SARS-CoV-2 cases, and then determine how this bias may affect sample size calculations. We then extend the framework and focus on genomic surveillance as an ongoing process, with sampling occurring periodically over time.

## METHODS

### Variant surveillance model

As discussed above, we aimed to characterize the factors that could affect the collection of SARS-CoV-2 samples and their selection for downstream processes such as sequencing. To do this, we developed a model that tracks how biological differences between variants, as well as logistical challenges in case and variant detection, may affect estimated variant frequency, given a true underlying frequency in a population. This model distinguishes all infections (*N*), from those that are detected (*D*), from those that ultimately produce a genome sequence (*G*) that can be used to identify the underlying variant.

We conceive of the model in two phases: 1) *infection detection*, which describes the joint biological and testing mechanisms that lead infections (*N*) to be detected (*D*) by a surveillance system (**Fig 3**, top row), and 2) *infection characterization*, which describes the selection of samples for genomic sequencing from high-quality detected infections (*H*) and identification of specific variants from the resulting high-quality sequences (**Fig 3**, bottom row). In this context, “high-quality detected infections” refers to pathogen-positive samples of high enough quality (e.g., by a metric such as cycle threshold value) that they will be selected for sequencing, while “high-quality sequences” refers to pathogen sequences that are complete enough to characterize the infection-causing variant.

**Figure 3.**
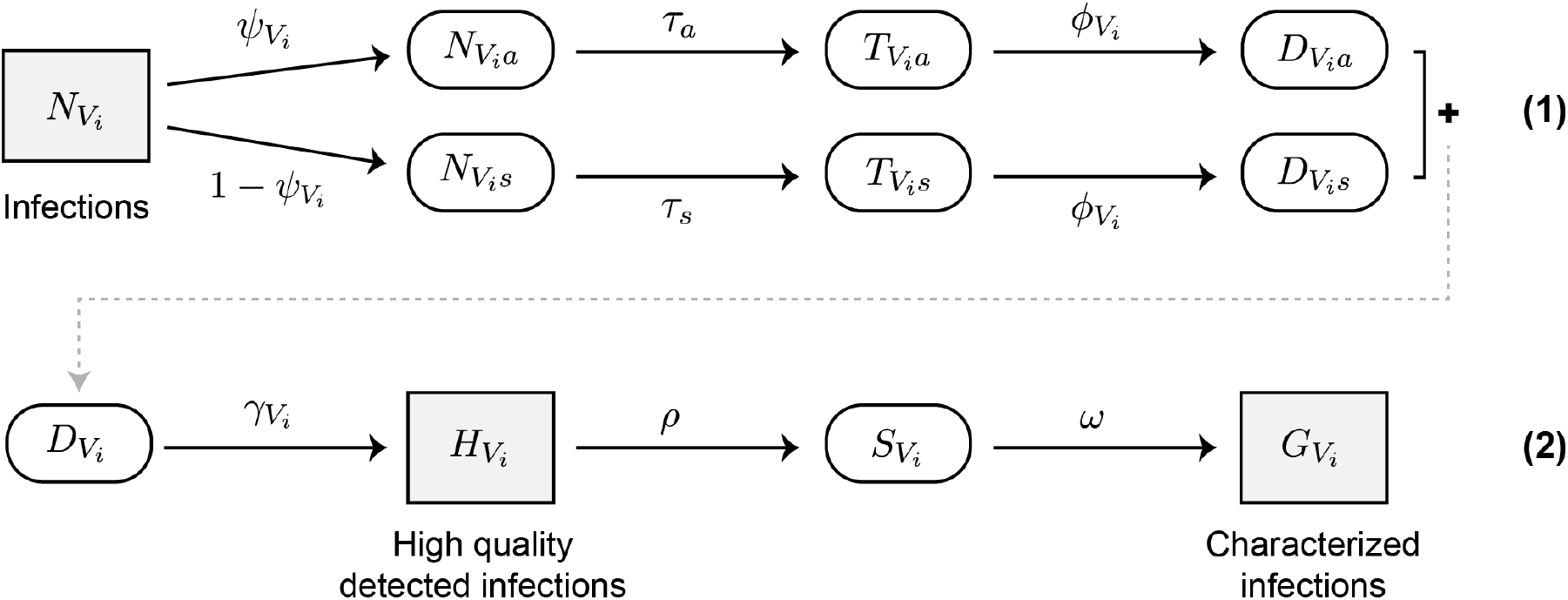
Schematic of variant surveillance model. (1) infection detection process; (2) infection characterization process. Parameters are defined in **Table 1**.

Model states are separated by transition parameters that model how biological differences between variants can affect factors such as testing rates, testing sensitivity, and sample quality (**Table 1**). Each of these parameters is specific to the relevant population scale and time period of sampling, e.g., the testing rate is the nationwide testing rate at the time of sampling if the model will be used to determine overall country-wide VOC prevalence but may be different if the method is to be applied at a smaller geographic scale, or in a setting with different testing practices or healthcare-seeking behaviors. The model is also generalizable for any number of variants of interest, where the VOC(s) are always compared to the remaining population. For example, the number of infections caused by a VOC *V*_1_ is tracked alongside the number of infections not caused by this variant, which we term *V*_2_. This setup allows us to track the proportion of infections caused by variant *i* at any given step, which is often the value of primary interest.

**Table 1.** Model states and parameters.

| Param | Description | Param | Description |
| --- | --- | --- | --- |
| $N$ | Total number of infections in population | $H_{V_i}$ | Number of detected, high quality, infections caused by variant $i$ |
| $P_{V_i}$ | Proportion of variant $i$ in population | $S_{V_i}$ | Number of detected, high quality, infections caused by variant $i$ that are selected for sequencing |
| $N_{V_i}$ | Number of infections caused by variant $i$ | $G_{V_i}$ | Number of high quality sequences from infections caused by variant $i$ |
| $N_{V_i,a}$ | Number of asymptomatic infections caused by variant $i$ | $\psi_{V_i}$ | Probability that an infection caused by variant $i$ is asymptomatic |
| $N_{V_i,s}$ | Number of symptomatic infections caused by variant $i$ | $\tau_x$ | Probability of testing, given type of infection ($x$: symptomatic or asymptomatic) |
| $T_{V_{ia}}$ | Number of tested asymptomatic infections caused by variant $i$ | $\phi_{V_i}$ | Probability that a tested infection caused by variant $i$ results in a positive test (sensitivity) |
| $T_{V_{is}}$ | Number of tested symptomatic infections caused by variant $i$ | $\gamma_{V_i}$ | Probability that a detected infection caused by variant $i$ meets some quality threshold |
| $D_{V_{ia}}$ | Number of detected asymptomatic infections caused by variant $i$ | $\rho$ | Probability that a sample is selected for sequencing |
| $D_{V_{is}}$ | Number of detected symptomatic infections caused by variant $i$ | $\omega$ | Probability that a sequenced sample produces a high quality genome |
| $D_{V_i}$ | Number of detected infections (symptomatic and asymptomatic) caused by variant $i$ | | |

Using this model, the number of high-quality detected infections attributable to a specific variant is as follows:

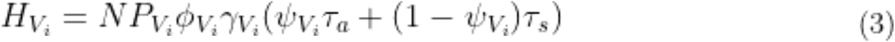

By calculating this quantity for each variant of interest and the remainder of the population, we can determine the prevalence of each VOC in the pool of high quality samples available for sequencing. Variation in these parameters between pathogen variants can be summarized in a single parameter, which we term the *coefficient of detection*:

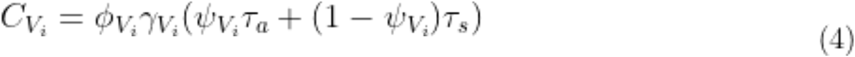

The value of this coefficient for each variant will determine the bias already present in *H*, the population from which we ultimately draw our sample (Fig S2). As shown below, only the ratio of variant coefficients (and not the raw value for each VOC) is necessary for sample size calculations, so it is not necessary to estimate every component parameter. When the detection parameters do not vary between variants, the ratio of the coefficients of infection will be one, and VOC prevalence in detected high quality infections will mirror prevalence in the overall population.

### Model assumptions

Although testing rate, testing sensitivity, and other parameters included in the core version of the model account for some of the major biases in infection detection and characterization, we do not include every potential source of bias, including spatial or temporal heterogeneity ^14^. Therefore, one of the critical underlying assumptions of our model is that sampling is homogeneous and representative across the relevant time period, geography of interest, and any other factors not explicitly included as model parameters.

The current version of the model also assumes that parameter values are static over time, and does not explicitly account for turnaround time (i.e., the amount of time between sample collection and variant characterization). Turnaround time can vary greatly across surveillance settings and countries ^3^, and therefore should be carefully considered when interpreting results based on sampling strategies suggested by the model. Any conclusions apply for the day on which samples were collected (rather than the date of sequencing, which could be days or weeks later) and, if samples were collected over a period of time, the user must make an additional assumption that the variant prevalence did not change significantly over that time period in order to correctly interpret the results.

Finally, we assume that all variants not explicitly tracked in application of the model are homogeneous in terms of testing rate, testing sensitivity, and other variant-specific parameters. In other words, if we are interested in tracking a variant *V*_1_, we assume all non-*V*_1_ infections of this virus (which we summarize as single variable, *V*_2_) have identical biological properties. Accounting for heterogeneity in *V*_2_ would require a three (or more) variant system, in which specific characteristics of variants *V*_1_, *V*_2_, etc. are compared to the remaining pathogen population, *V*_%_.

### Estimating the effects of pathogen properties on variant surveillance

We explored the effects of biological and logistical factors affecting variant detection—summarized in the coefficient of detection (**Equation 4**)—on the variant proportions observed in *H*, the pool of high quality detected infections from which to sample (**Equation 3**). We can calculate the multiplicative bias in the observed prevalence of a particular variant as a function of the underlying prevalences and coefficients of detection for all variants in the system:

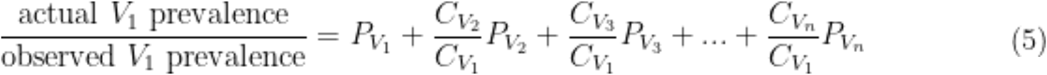

Where *n* is the total number of variants in the population (*n* ≥ 2).

Unsurprisingly, a larger differential between the coefficient of detection for *V*_1_ and the coefficients of detection for other variants in the system leads to more bias in the observed frequency. Additionally, the observed prevalence of *V*_1_ in *H* is more biased when 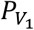 is smaller (**Fig 4A**).

**Figure 4.**
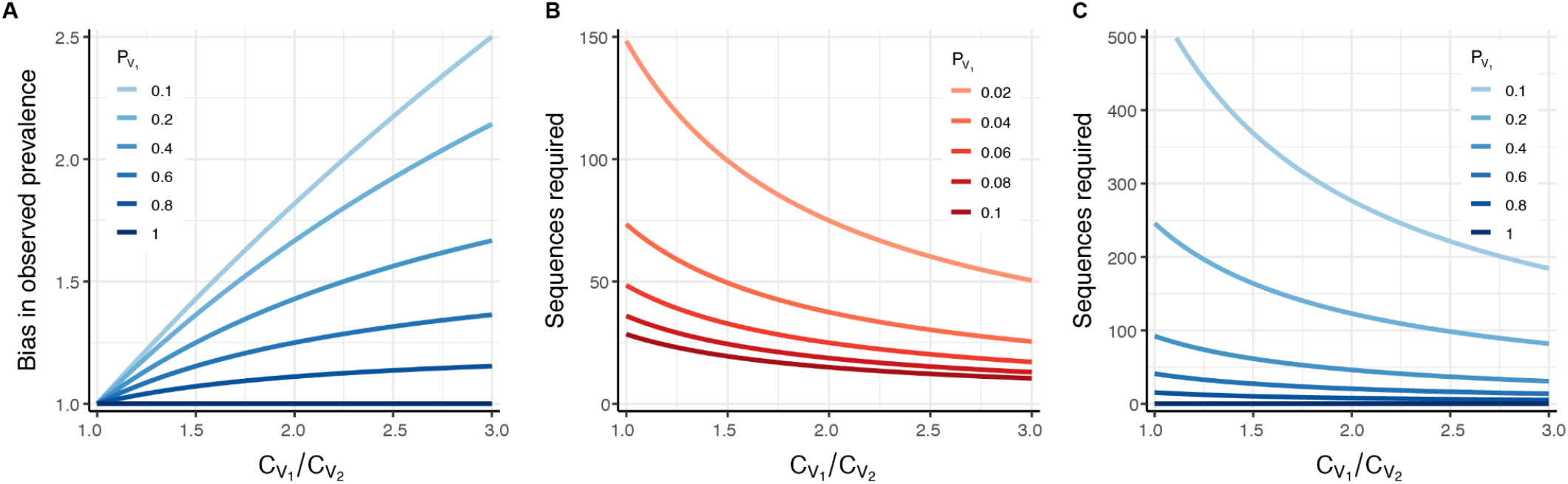
Exploring the effects of the coefficient of detection. (**A**) Multiplicative bias in the observed prevalence of variant *V*_*1*_ in *H*, the pool of high quality infected detections to sample from (bias: observed *V*_*1*_ prevalence divided by actual *V*_*1*_ prevalence). (**B**) Number of sequences required to detect at least one infection caused by *V*_*1*_ with 95% probability, for different *V*_*1*_ prevalence values and coefficient of detection ratios. (**C**) Number of sequences required to determine the prevalence of variants with a frequency of at least 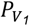 in the population, with 95% confidence and 25% precision. The prevalence calculated with these sequences will reflect the observed (biased) value, and will need to be corrected using **Equation 6**. All panels assume a two-variant system, where *V*_*1*_ is the variant of interest and *V*_*2*_ is the rest of the pathogen population. In (B) and (C), note that the number of samples selected for sequencing should exceed the number of sequences required if *ω* < *1*.

We can also calculate a correction factor *q* such that:

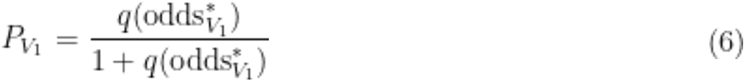

Where 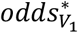 is the observed odds of the *V*_1_ prevalence in *H*. This equation allows for a direct conversion between the observed variant frequencies in the sampling pool 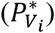 and the true frequency of *V*_1_ in the infected population 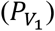. In a two-variant system (i.e., a system with one variant of interest compared to the rest of the population), 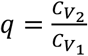 (see Supplemental Text 1 for derivation and correction factor values in larger systems).

### Implementation

We have implemented the method described above in a publicly available R package (*phylosamp*) and spreadsheet (Supplemental Data 1). Either can be used to calculate the required sample size in each of the three scenarios described below: cross-sectional sampling for variant detection, cross-sectional sampling for measuring variant prevalence, and periodic sampling for variant detection. The equations can be used both backwards and forwards— a user can input epidemiological and biological parameters and use them to determine the sample size needed to achieve the primary aim (detection or measuring prevalence) given a desired confidence level, or they can input an anticipated sample size and use the equations to calculate confidence in the results.

## RESULTS

### Sampling strategies for cross-sectional surveillance

In the following sections, we provide examples of how to calculate the appropriate sample size for surveillance given potential biases in observed variant frequencies. We also discuss how this bias—or, in some cases, enrichment—may make it easier to detect or measure the prevalence of certain variants. A complete, worked example of using our method to set up a surveillance program is provided in Supplementa**l Text 2**.

### Variant detection

Detecting the introduction of new variants into specific populations is a common goal during a pathogen outbreak. This requires identification of variants while they are still at low frequency in the population. For example, we may be interested in determining the minimum sample size needed to have a 95% chance of detecting a variant at 2% frequency in a specific population. If this variant is biologically and epidemiologically identical to the rest of the population, its frequency in the sampling pool (*H*) will reflect its frequency in the overall population. In this case, we can apply binomial sampling theory (**Equation 1**) to calculate the number of sequences needed (see **Fig S3** for validation of binomial sampling process):

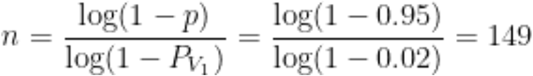

Most variants of interest, however, are not biologically and epidemiologically identical to the rest of the pathogen population. For example, variants can emerge that are more transmissible, such as the SARS-CoV-2 Delta variant ^1,15,16^. This increased transmissibility can be for a variety of reasons, such as higher viral loads in infected patients or more efficient entry into host cells, all of which require adjustment to these calculations. Here, we assume that a VOC is more transmissible specifically because it causes higher viral titers in infected patients, and that, based on current testing practices, this increased titer increases the testing sensitivity of the variant 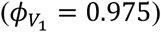 as compared to other circulating variants 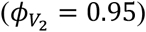. We also assume that detected infections caused by this VOC contain more virus and therefore have an increased probability 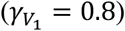 of meeting quality thresholds (e.g., Ct value cutoffs) using currently available sequencing technologies than other positive samples 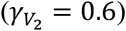. We assume that all other biological and surveillance parameters are the same between the VOC and the rest of the pathogen population, so we do not need to estimate their values in order to obtain the coefficient ratio needed for sample size calculations.

Using the parameters that differ, we can calculate the coefficient of detection ratio and use this to calculate the VOC frequency we expect to see in our sample. Rearranging **Equation 5**, we see that:

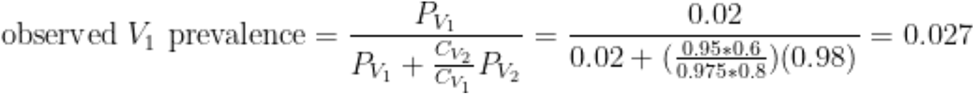

We then apply sampling theory as above, using the observed variant frequency (2.7%) as 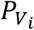. Because the variant is enriched in our population of detected infections, we find that only 109 sequences are needed to be 95% confident in detection of this variant (**Fig 4B**; see **Fig S4A** for sequence requirements for 50% confidence). Since not every sequenced sample produces a usable sequence, even after selecting for high quality samples (**Fig 3**), we assume a sequencing success rate of 80% for all variants (ω = 0.8), which means 137 samples should be selected for sequencing in order to obtain 109 complete genomes. The same procedure could be performed to determine the sample size needed to detect a more severe variant—or any variant that has some effect on pathogen detection— provided the ratio of coefficients of detection can be estimated. We note that all estimated parameters are specific to the population and time period of sampling, and would require modification for use in other settings, which could involve different testing practices, healthcare-seeking behaviors, or variant characterization technologies.

### Variant prevalence

After a variant is first detected, sequencing is often used to monitor its frequency in the population. Therefore, we assume that we are interested in calculating the minimum sample size needed to correctly (within 25% of the true value) determine the prevalence of a variant at >10% frequency in the population with 95% confidence. If this variant is biologically and epidemiologically identical to the rest of the population, its frequency in the sampling pool (*H*) will reflect its frequency in the population. In this case, we can apply existing theory (**Equation 2**) to calculate the number of sequences needed:

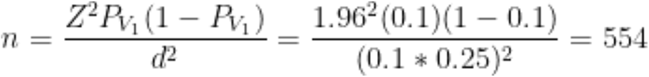

In this example, we calculate the sample size with the smallest prevalence (10%) we are interested in accurately measuring, since this requires the largest sample size. We do not apply any sort of finite population size correction ^8^, though this could decrease the sample size needed for prevalence estimation.

If a variant of interest has differing biological or epidemiological properties, we must adjust our calculations to account for the likely over or underrepresentation of this variant in the sampling pool. A more severe variant, for example, may decrease the proportion of infected individuals who are asymptomatic 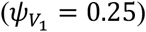, compared to the rest of the population 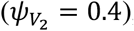, but may have a limited effect on the other biological and surveillance parameters. A difference in the asymptomatic rate only biases the observed variant frequency if testing rates are different for symptomatic and asymptomatic infections (see **Equation 4**), so in this example we assume that symptomatic infections are tested at a higher frequency (*τ*_*s*_ = 0.3) than asymptomatic infections (*τ*_*a*_ = 0.05). We again assume all other parameters are equal between variants (and thus do not provide a value for them) and use **Equations 4 and 5** to calculate the variant frequency we expect in our sample, assuming a true underlying frequency of 10%:

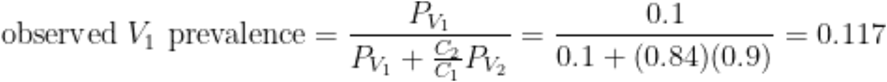

We then apply sampling theory as above, using 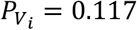. We find that the enrichment of VOC samples among detected infections means fewer sequences (*n* = 465; which would require sequencing 580 samples assuming an 80% sequencing success rate) are needed to achieve the desired precision in our estimate of variant prevalence (**Fig 4C**; see **Fig S4B-C** for sequence requirements with different confidence and precision values). However, it is important to keep in mind that, even if 465 sequences are successfully generated, the variant prevalence estimated from these data will be the *observed variant prevalence* and not the true population prevalence of the variant. In other words, even sequencing every sample available will not overcome the biases in the sampling pool itself. **Equation 6** must be applied to estimate the true variant prevalence from any observed value.

### Sampling strategies for ongoing surveillance

During an infectious disease outbreak, variant detection and monitoring are ongoing processes. Sampling over time is needed to conduct critical surveillance tasks, such as monitoring for the introduction of new variants into the population. Determining the appropriate sample size for this task requires us to adapt the above approaches.

### Variant detection

Here, we assume that the same number of sequences are sampled at each time step, e.g., that sequencing batches prepared weekly or daily always include the same number of samples. Given this assumption, we can again use binomial sampling theory (**Equation 1**) to calculate the probability of detecting a VOC on or before time step *t*. The resulting equation takes the form of a survival function, as follows:

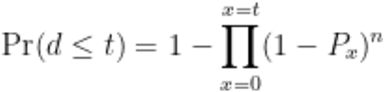

Where *Pr*(*d* ≤ *t*) is the probability of detection on or before time *t, n* is the sample size per unit time, and *P*_*x*_ is the prevalence of the variant of interest in the population at time *x*. After rearranging this equation to solve for the per-timestep sample size and approximating the product with a continuous function (see **Appendix** for full derivation), we obtain:

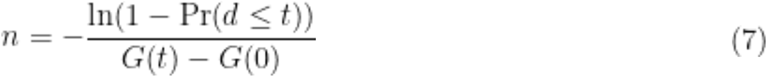

Where *G*(*t*) is the cumulative density of the function used to model variant growth over time. In other words, we can estimate the necessary sample size per time step, provided we can approximate how the variant prevalence is changing over time.

For example, let us assume that variant prevalence follows a logistic growth curve. Logistic growth is often ascribed to variants with a fitness advantage, such as the Alpha SARS-CoV-2 variant ^17^, though in this section we will assume that the variant of interest does not affect any of the parameters that go into calculating the coefficient of detection (we will relax this assumption in the following section). We assume there was a single introduction of this variant into a population of 10,000 infected individuals and that the growth rate is approximately 0.1 per day ^18^. Now, we can use **Equation 7** to calculate the per-day sample size needed to ensure detection (with 95% probability) of this Alpha-like variant within 30 days of its initial emergence:

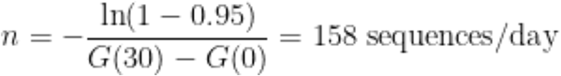

In other words, generating 158 * 7 = 1,106 sequences per week (assuming sequences are well-distributed throughout the week) ensures a 95% probability of detection of this variant within 30 days of initial introduction. Over 1,000 sequences per week may be an unmanageable number, but it is important to note that, given the assumptions of a single introduction and a growth rate of 0.1 per day, the variant will only have a prevalence of 0.2% by day 30. It may be more realistic to use these logistic growth assumptions to estimate when prevalence will surpass a specific, more detectable value (e.g., with these assumptions, prevalence will surpass 1% on day 47) and then to determine the sample size needed to ensure detection before the VOC reaches that prevalence. Inserting 47 days into **Equation 7** shows us that a much more manageable 196 sequences per week are needed to detect the variant before it surpasses 1% prevalence in the population of interest.

Once again, it is also important to consider the sequencing success rate (*ω*) when calculating the number of samples that should be selected for sequencing. To generate 196 high quality sequences per week with an 80% success rate, 245 samples will need to be selected for sequencing.

### Variant detection with a biased sample

As discussed above, VOC prevalence may be enriched in the sampling pool, meaning that fewer sequences may be needed for confident detection of the variant. Using **Equation 5**, we can calculate the observed variant frequency at each time step given a growth rate and starting variant prevalence (e.g., 1 introduction into an infected population of 10,000) as follows:

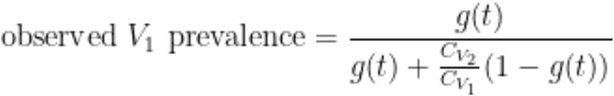

Where 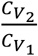 represents the relative coefficients of detection between the general pathogen population 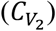 and the variant of interest 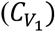. Furthermore, *g*(*t*) is the probability density function used to model variant growth over time. From this, we can calculate the cumulative density function of the *observed* variant prevalence distribution, *G*\* (*t*), and use this approximation in our sample size calculation.

If we assume that the Alpha-like VOC described above results in a coefficient of detection ratio of 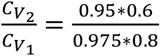 (see ‘Variant Detection’ in ‘Sampling Strategies for Cross-Sectional Surveillance’), the sample size needed to ensure a 95% probability of detection by the time the VOC prevalence surpasses 1% in the population is:

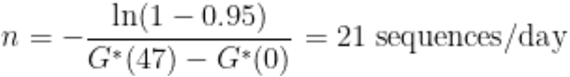

Where we again assume an initial prevalence of 1 in 10,000 and a growth rate of 0.1 per day (see **Appendix** for full derivation). As expected, the enrichment of the VOC in the sampling pool decreases the number of sequences needed for detection. **Figure 5** (and **Fig S5**) lays out the relationship between detection probability, sample size, and variant growth, and can be used to evaluate the marginal costs and benefits of changing the number of samples selected for sequencing. When using these curves to aid in the design of surveillance systems that take maximum advantage of available resources, it is important to remember that the coefficient of detection may not remain static over time. For example, testing infrastructure and practices changed multiple times over the course of the COVID-19 pandemic (e.g., testing increased with increased availability of approved diagnostic kits, then decreased dramatically as at-home testing became available and encouraged), which could alter testing rates, testing sensitivity, etc.—key inputs in the current version of our model. Accordingly, calculations should be repeated and sampling adjusted each time there is a significant shift in estimated parameter values due to behavioral changes, updates to variant characterization technology, or the emergence of new variants with distinct biological properties.

**Figure 5.**
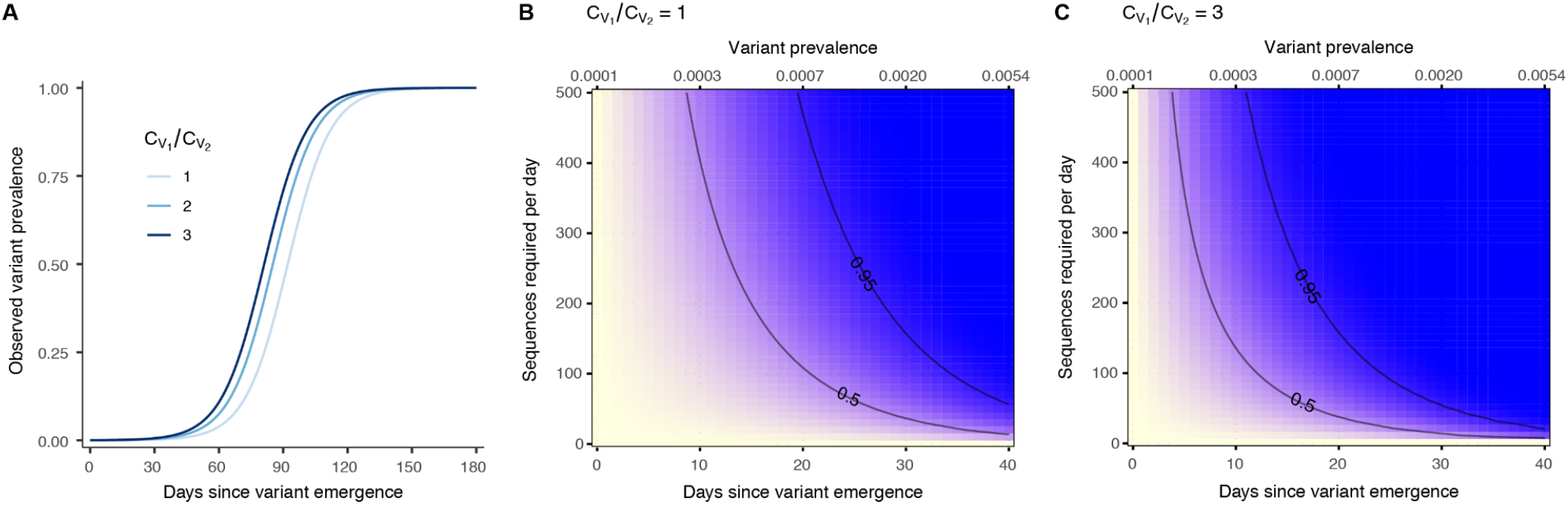
Sample size required for detection of a variant growing in prevalence. For a variant whose prevalence increases following a logistic curve with growth rate = 0.1 per day and starting value = 1/10000: (**A**) Observed variant prevalence over time given different coefficient-of-detection ratios. (**B-C**) Probability of detecting at least one infection caused by *V*_*1*_ (yellow = 0% probability; blue = 100% probability) on or before a specific day (bottom x-axis) or desired prevalence (top x-axis), given per-day sample size and specified coefficient of detection ratio. Note that the desired prevalence (top x-axis) is the actual variant prevalence in the population and that the number of samples selected for sequencing should exceed the number of sequences required if *ω* < *1*. 50% and 95% probability of detection contours are indicated.

### Variant prevalence

When monitoring variant prevalence is the primary goal, ongoing sampling strategies can improve prevalence estimates by leveraging multiple samplings of the infected population. In other words, we can view repeated samples (i.e., sequencing batches, and the VOC prevalence estimates obtained from these batches) as correlated data, where estimates from recent past data points are weighted according to their distance from the present time. Given the same parameters, this type of approach lowers the number of sequences that must be generated at each time step as compared to a cross-sectional sampling approach, while placing greater weight on recent samples that might better reflect current prevalence.

Specifically, in the cross-sectional monitoring framework, the prevalence estimate obtained from sequenced samples most likely represents the VOC prevalence in the middle of the period from which samples were collected, assuming consistent sampling over time. This estimate, while potentially useful, will be problematic if prevalence rapidly changes during that time period. Therefore, it may be useful to design a sampling scheme where samples are collected at fixed (and more narrow) time intervals, so that estimates present a more real-time picture of variant prevalence. Furthermore, information from recent time points can be used to inform the current estimate using a rolling window approach, with larger weight given to more recent data.

If we assume a constant sample size (i.e., number of sequences characterized) at each time step and a constant variance in prevalence estimates across time steps, we can show that the effective sample size across all time steps is (see **Appendix** for full derivation):

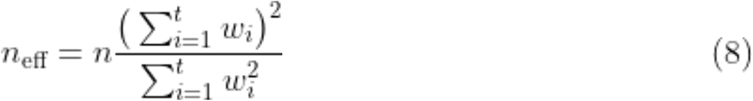

Where *n* is the per-timestep sample size, *t* is the total number of time points used in our prevalence calculation, and *W*_*i*_ is the weight given to a particular time point *i*. This is an application of Kish’s equation ^19^, and can be used to calculate the effective sample size for any particular weighting scheme. Since the effective sample size is the required sample size in **Equation 2**, we can easily rearrange **Equation 8** above to calculate the required per-timestep sample size needed to estimate prevalence with a desired confidence and precision:

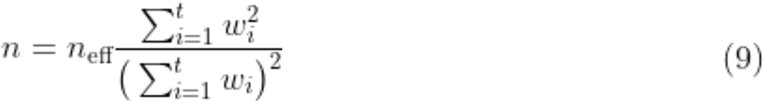

As in the example used in the cross-sectional section above, let us assume that we are interested in calculating the minimum sample size needed to correctly (within 25% of the true value) determine the prevalence of a variant at >10% frequency in the population with 95% confidence. **Equation 2** shows that this will require 554 sequences. Instead of collecting all of these sequences at once, we will sample infections from the population weekly and calculate prevalence using the following weighting scheme:

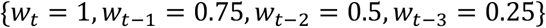

Using **Equation 9**, we can calculate the required number of sequences needed per week as follows:

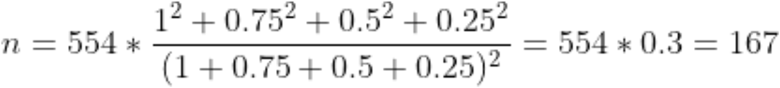

In other words, generating 167 sequences each week and applying the weighting scheme described above to calculate prevalence will generate an estimate with the desired confidence and precision. This requires substantially fewer resources than generating 554 sequences each week, while resulting in an estimated prevalence that may be more reflective of the current prevalence than spreading the 554 sequences equally across samples collected from infections spanning the four-week time period in this example. The choice of weighting scheme will of course affect the relevance of the prevalence estimate and we direct the reader to the extensive literature on selecting appropriate weighting schemes ^20–23^, a discussion of which is outside the scope of this manuscript.

### Variant prevalence with a biased sample

This methodology can also be used if the sampling pool itself is biased. In this case, we use **Equations 4 and 5** to calculate the observed prevalence and use this value in **Equation 2**, as described above. The required sample size determined from **Equation 2** (the effective sample size) can then be used to calculate the number of sequences required per timestep to achieve desired confidence and precision in variant prevalence estimates.

## DISCUSSION

Designing a pathogen surveillance system in a particular population must begin with identifying the primary purpose or key questions to be answered. For example, surveillance strategies will be different when the goal is early detection of a newly introduced VOC versus when the goal is measuring the prevalence of an existing variant^12^. In either case, there are a myriad of factors that influence which infections are ultimately sequenced. Here we present a framework for thinking about these factors, and we show that their effects can be summarized in a single number, the coefficient of detection. This coefficient characterizes how biological and logistical factors can lead to VOC enrichment (or depletion) in a sample—leading to earlier (or later) detection—while also biasing measurement of the true underlying VOC prevalence. Depending on the purpose of surveillance, it will be important to account for these effects in sample size calculations and subsequent reporting of results. The work presented here aims to provide an accessible set of methods for doing so, and a general approach that can be extended to other settings and study designs. In addition to providing statistically-grounded guidance for sampling design, our framework can be applied retrospectively to evaluate previously reported results.

A perceived barrier to using the approach outlined here may be lack of knowledge of the exact parameters that are summarized by the coefficient of detection. However, it is not necessary to know individual parameter values when using our framework as long as we can approximate their ratio, since all calculations rely solely on the ratio of coefficients of detection. Parameters that have the same value across variants need not be specified at all. Although decreasing the number of parameters that need to be specified makes the framework easier to use, there is still value in breaking down the process of surveillance into its component parts. For instance, it may be difficult to estimate a single pathogen testing rate in settings without consistent testing of asymptomatic individuals (e.g., hospitals or settings with limited testing capacity). But if the asymptomatic and symptomatic testing rates are separated into two parameters, we can assume the asymptomatic testing rate is negligible (or at least similar between variants) and focus on the symptomatic testing rate, which may be easier to quantify.

In considering the full process from infection detection to variant characterization, we have aimed to make our framework flexible enough to handle situations not explicitly discussed above. Although most of the examples presented in this manuscript focus on a two-variant model, the framework is set up to allow for exploration of multiple variants simultaneously (see **Appendix**). Further, while we focus on detecting variants undergoing logistic growth, the sample size needed to detect a variant can be calculated for any growth function as long as the functional form and underlying parameters can be approximated. Additionally, while we have tried to identify the key processes that affect pathogen detection, the coefficient of detection could be modified to incorporate other parameters that differ between variants, or to include factors that may affect which sequences produce complete genomes (i.e., factors that affect the variant characterization process shown in the bottom part of **Fig 3**). For example, we could allow the sequencing success rate (*ω*) to differ between variants (e.g., due to differences in primer binding when using PCR-based methods for variant characterization or amplification prior to sequencing), despite the use of an initial sample quality filter (*γ*). Finally, the framework could be extended to any pathogen for which there is some method (e.g., variant-specific PCR assays) to differentiate pathogen lineages with potentially different epidemiological or biological processes.

While this manuscript provides guidance for sample size calculations in a number of contexts, it is far from comprehensive. However, we hope that the methods presented here provide a good starting point for more sophisticated approaches or those that are more precisely tailored to a specific context. For instance, additional work on incorporating previous measurements in variant prevalence estimation is needed; our calculations assume that the variance in prevalence estimates remains constant across time points, which may not be the case when variant prevalence is changing rapidly. Additionally, when multiple variants are present in the population, accurate prevalence estimation of one VOC necessarily constrains the potential prevalence values of another VOC; expanding the framework to co-estimate prevalences for multiple VOCs may make it possible to leverage this interdependence and further reduce the sample sizes required for accurate monitoring.

When designing surveillance systems based on this framework, it is important to remember that infection and sampling processes can be heterogeneous in ways not captured by our model. Future work could consider the effects of spatial heterogeneity on transmission and sampling, the impact of time-varying model parameters in the ongoing surveillance context, or could extend the framework to metapopulations. Given the current framework and assumption of homogeneity, samples selected for sequencing should be selected as randomly as possible (in relation to unmodeled factors), or selected in a way that maximizes the geographic and temporal distribution of sequences.

The intensive monitoring of SARS-CoV-2 genomic variation during the pandemic has revealed the importance of characterizing and monitoring specific pathogen variants. As sequencing technologies become more accessible and central to our understanding of established and emerging pathogens, it is important that we improve the rigor with which we design studies using this data. Sophisticated modeling approaches have been invaluable in improving how we collect and interpret pathogen genomic information, but most are neither nimble nor accessible enough to be widely used during a crisis. Similarly, ad-hoc approaches or classical study designs may not lead to the optimal allocation of resources. Here we have attempted to lay out a framework that is widely accessible yet still accounts for many of the factors that uniquely impact pathogen genomic studies and surveillance programs. As the SARS-CoV-2 pandemic continues and new infectious threats arise, we hope this approach will help better guide the collection of data that has proven critical to the pandemic response and serve as a starting point for further methodological innovation.

## Supporting information

Supplemental Data 1 (Variant Sampling Workbook)

## Data Availability

Code to produce figures is available at: https://github.com/HopkinsIDD/VOCsamplesize.

## ACKNOWLEDGMENTS

We thank Edyth Parker for her insightful comments on the manuscript. Funding was provided by Bill and Melinda Gates Foundation INV-025321 (S.W.) and OPP1195157 (S.W. and J.L.).

## AUTHOR CONTRIBUTIONS

Conceptualization, S.W., E.C.L., and J.L.; Methodology, S.W., E.C.L., and J.L.; Software, S.W. and E.C.L.; Formal Analysis, S.W.; Resources, J.L.; Writing – Original Draft, S.W., B.L.D., and E.C.L.; Writing – Review & Editing, S.W., E.C.L., B.L.D., and J.L.; Visualization, S.W. and E.C.L.; Supervision, J.L.; Funding Acquisition, S.W. and J.L.

## DATA AND CODE AVAILABILITY

All code, including code to generate figures, is publicly available here: https://github.com/HopkinsIDD/VOCsamplesize.

## REFERENCES

1. Tracking SARS-CoV-2 variants https://www.who.int/en/activities/tracking-SARS-CoV-2-variants/.

2. Public Health England Investigation of novel SARS-CoV-2 variant: Variant of Concern 202012/01.

3. Tegally, H., Wilkinson, E., Giovanetti, M., Iranzadeh, A., Fonseca, V., Giandhari, J., Doolabh, D., Pillay, S., San, E.J., Msomi, N., et al. (2020). Emergence and rapid spread of a new severe acute respiratory syndrome-related coronavirus 2 (SARS-CoV-2) lineage with multiple spike mutations in South Africa. bioRxiv. 10.1101/2020.12.21.20248640.

4. Faria, N.R., Mellan, T.A., Whittaker, C., Claro, I.M., Candido, D. da S., Mishra, S., Crispim, M.A.E., Sales, F.C.S., Hawryluk, I., McCrone, J.T., et al. (2021). Genomics and epidemiology of the P.1 SARS-CoV-2 lineage in Manaus, Brazil. Science. 10.1126/science.abh2644.

5. Classification of Omicron (B.1.1.529): SARS-CoV-2 Variant of Concern https://www.who.int/news/item/26-11-2021-classification-of-omicron-(b.1.1.529)-sars-cov-2-variant-of-concern.

6. Bushman, M., Kahn, R., Taylor, B.P., Lipsitch, M., and Hanage, W.P. (2021). Population impact of SARS-CoV-2 variants with enhanced transmissibility and/or partial immune escape. Cell. 10.1016/j.cell.2021.11.026.

7. Wohl, S., Giles, J.R., and Lessler, J. (2021). Sample size calculation for phylogenetic case linkage. PLoS Comput. Biol. 17, e1009182.

8. European Centre for Disease Prevention and Control (18-January-2021). Sequencing of SARS-CoV-2: first update.

9. Vavrek, D., Speroni, L., Curnow, K.J., Oberholzer, M., Moeder, V., and Febbo, P.G. (2021). Genomic surveillance at scale is required to detect newly emerging strains at an early timepoint. bioRxiv. 10.1101/2021.01.12.21249613.

10. The University of Texas COVID-19 Modeling Consortium Sample Size Calculator Detecting COVID-19 Variants. Variant Detection Calculator. https://covid-19.tacc.utexas.edu/dashboards/variants/.

11. Brito, A.F., Semenova, E., Dudas, G., Hassler, G.W., Kalinich, C.C., Kraemer, M.U.G., Hill, S.C., Danish Covid-19 Genome Consortium, Sabino, E.C., Pybus, O.G., et al. (2021). Global disparities in SARS-CoV-2 genomic surveillance. medRxiv. 10.1101/2021.08.21.21262393.

12. European Centre for Disease Prevention and Control (2021). Guidance for representative and targeted genomic SARS-CoV-2 monitoring.

13. Wayne W. Daniel, C.L.C. (2018). Biostatistics: A Foundation for Analysis in the Health Sciences, 11th Edition (Wiley).

14. Kraemer, M.U.G., Hill, V., Ruis, C., Dellicour, S., Bajaj, S., McCrone, J.T., Baele, G., Parag, K.V., Battle, A.L., Gutierrez, B., et al. (2021). Spatiotemporal invasion dynamics of SARS-CoV-2 lineage B.1.1.7 emergence. Science 373, 889–895.

15. Liu, Y., and Rocklöv, J. (2021). The reproductive number of the Delta variant of SARS-CoV-2 is far higher compared to the ancestral SARS-CoV-2 virus. J. Travel Med. 28. 10.1093/jtm/taab124.

16. Challen, R., Dyson, L., Overton, C.E., Guzman-Rincon, L.M., Hill, E.M., Stage, H.B., Brooks-Pollock, E., Pellis, L., Scarabel, F., Pascall, D.J., et al. (2021). Early epidemiological signatures of novel SARS-CoV-2 variants: establishment of B.1.617.2 in England. bioRxiv. 10.1101/2021.06.05.21258365.

17. Volz, E., Mishra, S., Chand, M., Barrett, J.C., Johnson, R., Geidelberg, L., Hinsley, W.R., Laydon, D.J., Dabrera, G., O’Toole, Á., et al. (2021). Assessing transmissibility of SARS-CoV-2 lineage B.1.1.7 in England. Nature 593, 266–269.

18. Davies, N.G., Abbott, S., Barnard, R.C., Jarvis, C.I., Kucharski, A.J., Munday, J.D., Pearson, C.A.B., Russell, T.W., Tully, D.C., Washburne, A.D., et al. (2021). Estimated transmissibility and impact of SARS-CoV-2 lineage B.1.1.7 in England. Science 372. 10.1126/science.abg3055.

19. Kish, L. (1965). Survey Sampling. Biometrical Journal 10. 10.1002/bimj.19680100122.

20. Zhang, X., and Wang, J.-L. (2018). Optimal weighting schemes for longitudinal and functional data. Stat. Probab. Lett. 138, 165–170.

21. Zhang, X., and Wang, J.-L. (2016). From sparse to dense functional data and beyond. aos 44, 2281–2321.

22. Tamiz, M., and Azmi, R. (2016). An investigation of various weighting schemes for portfolios. J. Bus. & Fin. Aff. 5. 10.4172/2167-0234.1000203.

23. Kalton, G., and Flores-Cervantes, I. (2003). Weighting methods. Journal of Official Statistics 19, 81–97.

