## Supplemental Information for "Sample Size Calculations for Variant Surveillance in the Presence of Biological and Systematic Biases"

**1 Supplemental Figures** **2**

**4 Supplemental Data 1: Variant Sampling Workbook (.xlsx)**

### Supplemental Figures

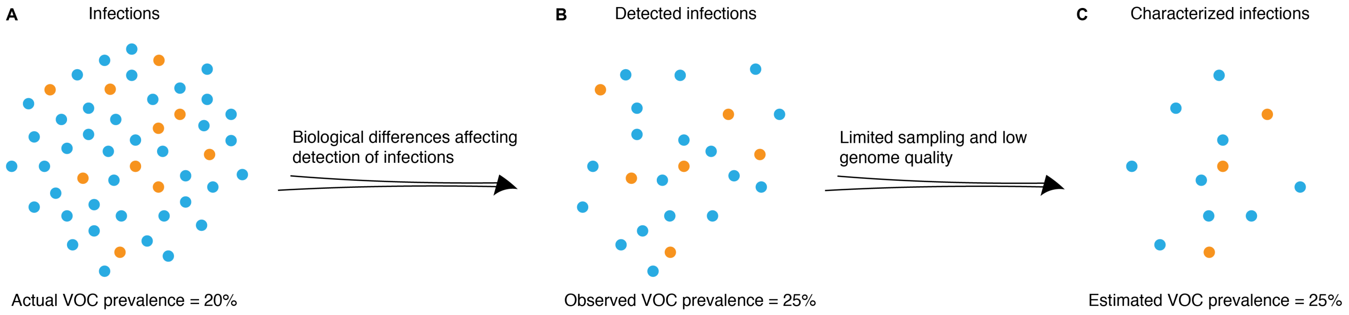

**Figure S1. Factors resulting in enrichment of observed variant prevalence.** VOC prevalence in (**A**) total population, (**B**) pool of detected infections, and (**C**) characterized infections (identified as a particular variant by sequencing or another technology). Biological differences between variants can lead to enrichment of VOC in observed variant proportions. Orange = infections caused by VOC (variant of concern); blue = infections caused by other variants of the same pathogen.

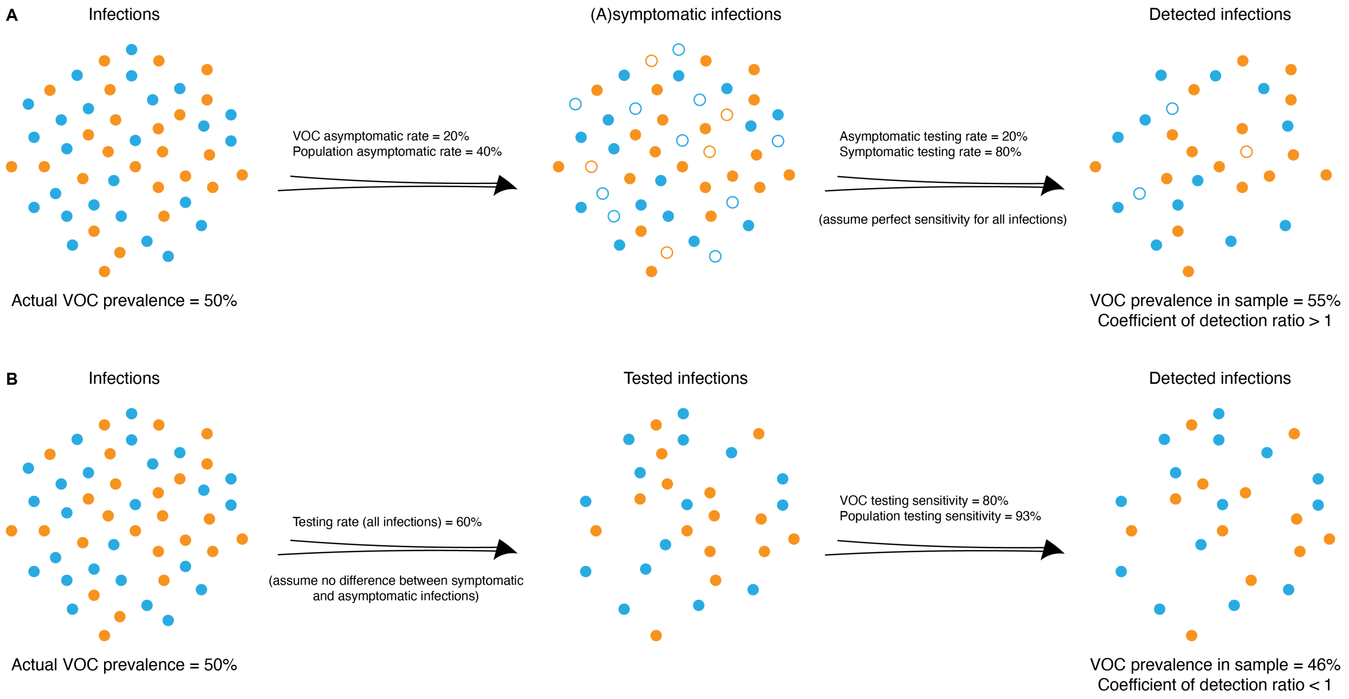

**Figure S2. Parameters affecting the coefficient of detection.** Factors affecting the variant of concern (VOC) prevalence in the sample of detected infections. In (**A**), a VOC (orange) causes more symptomatic infections (lower asymptomatic rate) than the rest of the pathogen population (blue). Assuming the testing probability is higher for individuals presenting symptoms than asymptomatic individuals, this leads to an artificial enrichment of the VOC in the pool of detected infections, since individuals infected with this variant are more likely to be tested and therefore sampled. Enrichment of a variant in a sample corresponds to a coefficient of detection ratio ($\frac{C_{V_1}}{C_{V_2}}$) that is greater than one. In (**B**), a VOC (orange) results in lower sensitivity of diagnostic tests, perhaps because it causes a lower viral load than the pathogen variant infecting the rest of the population (blue). In this case, the pool of detected infections is enriched for non-VOC samples, corresponding to a coefficient of detection ratio ($\frac{C_{V_1}}{C_{V_2}}$) of less than one.

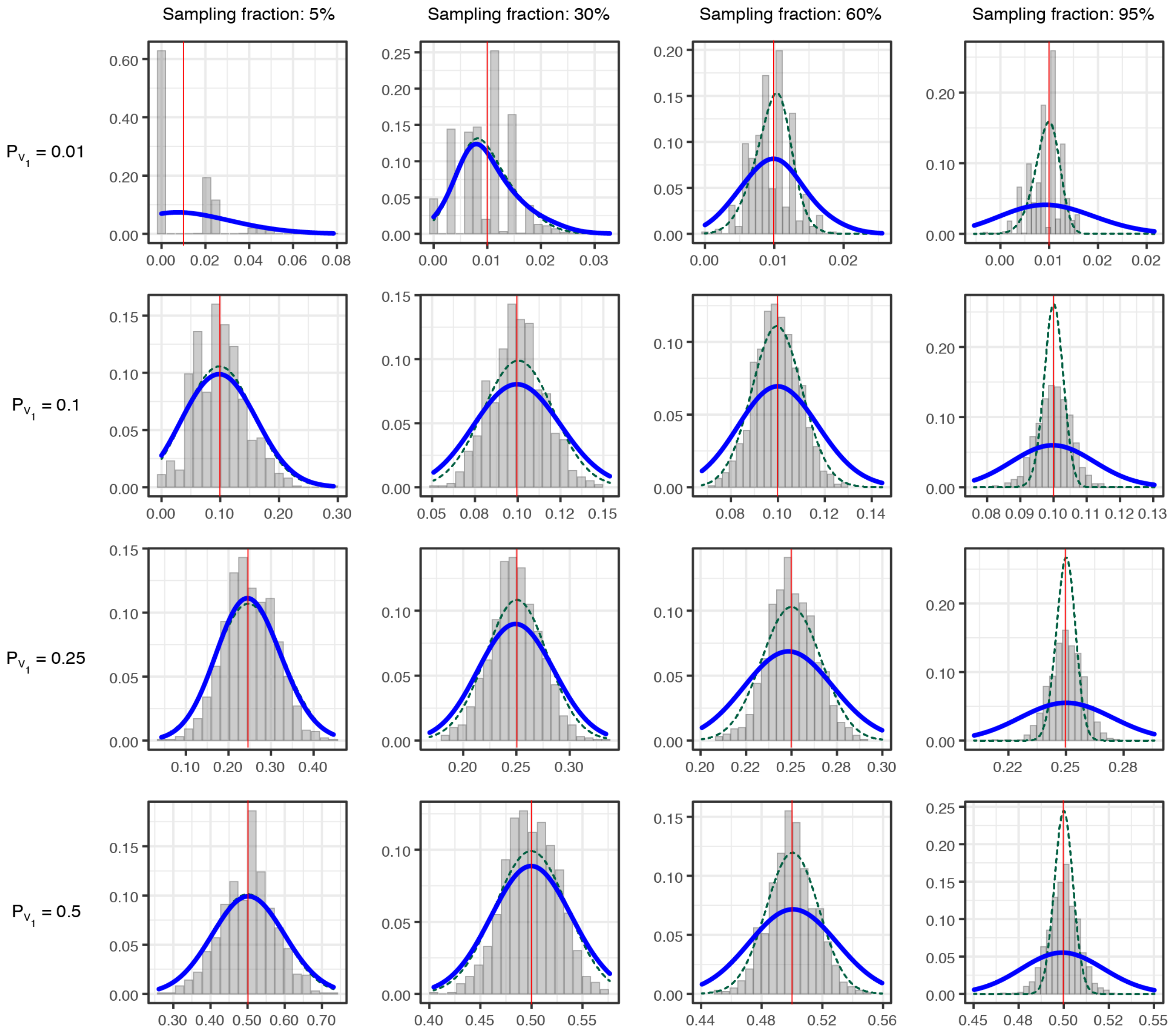

**Figure S3. Validation of the binomial sampling approximation.** Variant prevalence estimates (x-axis) from 1,000 simulations of our model for each combination of variant prevalence ($P_{V_1}$) (rows) and sampling proportion (columns). Simulations each assume 10,000 infected individuals in the population and $C_{V_1} = C_{V_2} = 0.114$, resulting in 1,140 high quality detected infections ($H$) from which to sample. In each simulation, =0.8 and infections that progress between model states are selected stochastically using a binomial process. Blue lines = binomial distribution (i.e., sampling with replacement, an approximation of the simulated sampling process) given stated sampling fraction (from $H$) and variant prevalence; green dotted lines = hypergeometric distribution (i.e., sampling without replacement, the exact sampling process) given stated sampling fraction and variant prevalence; red vertical lines = marker of input variant frequency. The binomial distribution approximates the simulated process well, except when nearly all detected infections are sampled (as expected) or the variant prevalence is very low.

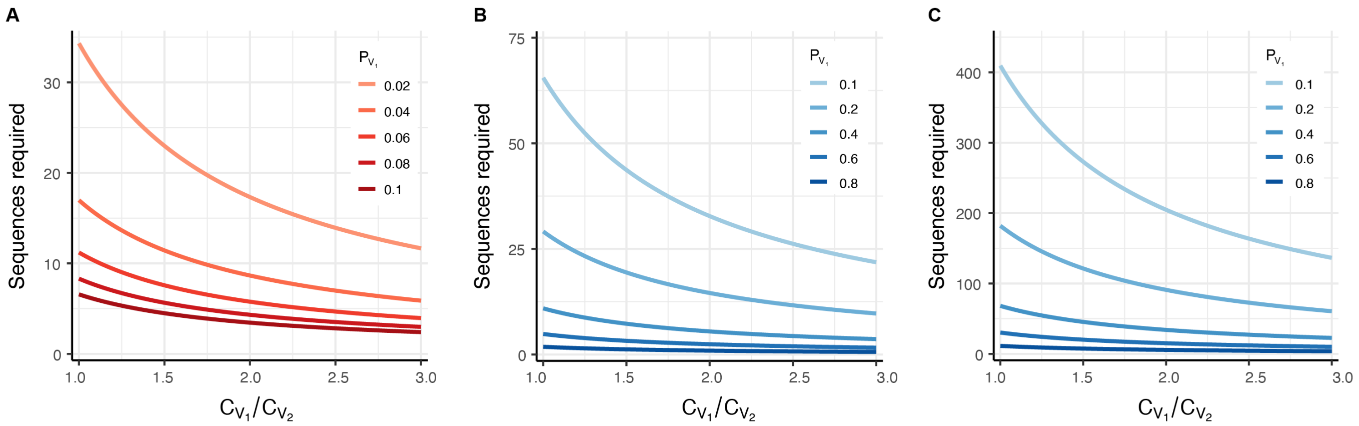

**Figure S4. Sample size needed for variant detection and prevalence estimation with 50% confidence.** (**A**) Number of sequences required to detect at least one infection caused by $V_1$ with 50% probability, for different $V_1$ prevalence values and coefficient of detection ratios. (**B**) Number of sequences required to determine the prevalence of variants with a frequency of at least $P_{V_1}$ in the population, with 50% confidence and 25% precision. (**C**) Same as (B), but with 50% confidence and 10% precision. The calculated prevalence in (B) and (C) will reflect the observed (biased) value, and will need to be corrected using (**Equation 6**). These figures assumes a two-variant system, where $V_1$ is the variant of interest and $V_2$ is the rest of the pathogen population. In all panels, note that the number of samples selected for sequencing should exceed the number of sequences required if $\omega < 1$.

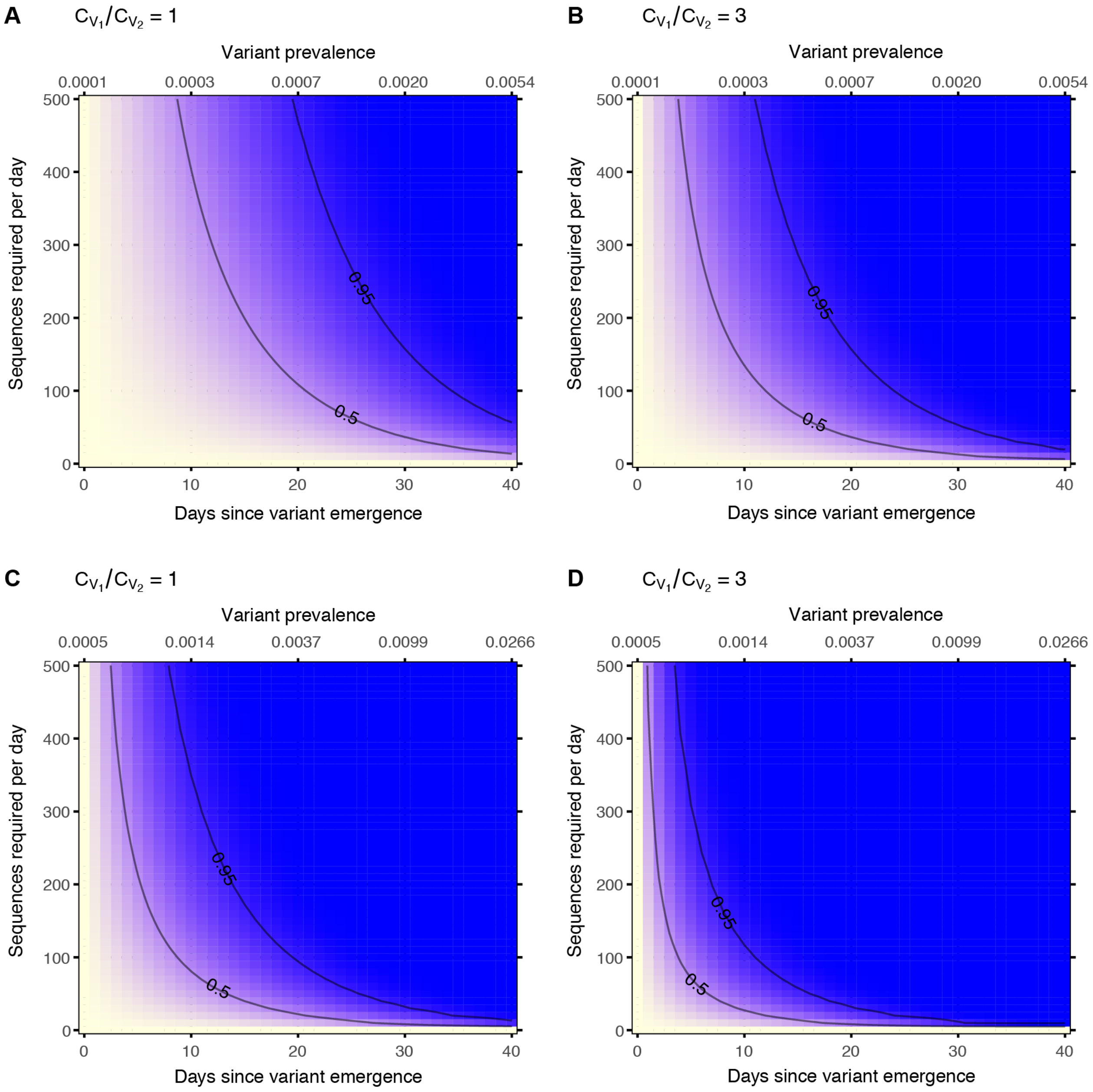

**Figure S5. Sample size needed for detection of a variant growing in prevalence.** Probability of detecting at least one infection caused by $V_1$ (yellow = 0% probability; blue = 100% probability) on or before a specific day (bottom x-axis) or desired prevalence (top x-axis), given per-day sample size, specified coefficient of detection ratio, and the following growth rate and initial variant prevalence values: (**A-B**) Growth rate = 0.05, initial prevalence = 1/10000. (**C-D**) Growth rate = 0.1, initial prevalence = 5/10000. Note that the desired prevalence (top x-axis) is the actual variant prevalence in the population and that the number of samples selected for sequencing should exceed the number of sequences required if $\omega < 1$. 50% and 95% probability of detection contours are indicated on all panels.

### Supplemental Text 1: Derivations

#### 1 Calculating variant prevalences from an observed sample

Given the coefficient of detection ($C_{V_i}$) for each variant in the pathogen population, we can calculate the actual prevalence of each variant ($P_{V_i}$) from what we observed in the pool of high quality detected infections ($H$). In the sections below, we use a property of odds ratios to calculate a correction factor $q$ that allows for this conversion.

##### 1.1 Correction factor in a 2-variant system

In a two-variant system (i.e., a system with one variant of interest, $V_i$, that is compared to the rest of the population, $V_2$), the odds of $V_1$ is:

$$\text{odds}_{V_1} = \frac{P_{V_1}}{P_{V_2}} = \frac{P_{V_1}}{1 - P_{V_1}}$$

Similarly, the observed odds (the odds of $P_{V_1}$ in $H$) is:

$$\text{odds}^*_{V_1} = \frac{P^*_{V_1}}{P^*_{V_2}} = \frac{\frac{C_{V_1}P_{V_1}N}{C_{V_1}P_{V_1}N + C_{V_2}P_{V_2}N}}{\frac{C_{V_2}P_{V_2}N}{C_{V_1}P_{V_1}N + C_{V_2}P_{V_2}N}} = \frac{C_{V_1}P_{V_1}}{C_{V_2}P_{V_2}} = \frac{C_{V_1}P_{V_1}}{C_{V_2}(1 - P_{V_1})}$$

We define a bias factor, $q$, such that:

$$\text{odds}_{V_1} = q \times \text{odds}^*_{V_1}$$

$$\frac{P_{V_1}}{1 - P_{V_1}} = q \times \frac{C_{V_1}P_{V_1}}{C_{V_2}(1 - P_{V_1})}$$

If we solve the above equation for $q$, we obtain:

$$q = \frac{C_{V_2}}{C_{V_1}}$$

.

Because we know that: $P_{V_i} = \frac{\text{odds}_{V_i}}{1 + \text{odds}_{V_i}}$, we can use the correction factor $q$ to easily calculate the true proportion of any variant $i$ in a population from its observed proportion in the sample $H$:

$$P_{V_i} = \frac{\text{odds}_{V_i}}{1 + \text{odds}_{V_i}} = \frac{q(\text{odds}^*_{V_i})}{1 + q(\text{odds}^*_{V_i})}$$

##### 1.2 Correction factor in a 3-variant system

In a 3-variant system, the odds of $V_1$ are as follows:

$$\text{odds}^*_{V_1} = \frac{P^*_{V_1}}{P^*_{V_2} + P^*_{V_3}} = \frac{\frac{C_{V_1}P_{V_1}N}{C_{V_1}P_{V_1}N + C_{V_2}P_{V_2}N + C_{V_3}P_{V_3}N}}{\frac{C_{V_2}P_{V_2}N + C_{V_3}P_{V_3}N}{C_{V_1}P_{V_1}N + C_{V_2}P_{V_2}N + C_{V_3}P_{V_3}N}} = \frac{C_{V_1}P_{V_1}}{C_{V_2}P_{V_2} + C_{V_3}P_{V_3}}$$

Using this, we can calculate $q_{V_{1,123}}$, the correction factor between the true and observed odds when $V_1$ is the variant of interest in a 3-variant system with $V_1$, $V_2$,and $V_3$:

$$\frac{P_{V_1}}{1-P_{V_1}} = q \times \frac{C_{V_1}P_{V_1}}{C_{V_2}P_{V_2} + C_{V_3}P_{V_3}} = q \times \frac{C_{V_1}P_{V_1}}{(C_{V_2}P_{V_2} + C_{V_3}P_{V_3})(\frac{1-P_{V_1}}{1-P_{V_1}})}$$

$$= q\Big(\frac{P_{V_1}}{1-P_{V_1}}\Big)\frac{C_{V_1}}{\frac{C_{V_2}P_{V_2}}{1-P_{V_1}} + \frac{C_{V_3}P_{V_3}}{1-P_{V_1}}}$$

$$= q\Big(\frac{P_{V_1}}{1-P_{V_1}}\Big)\frac{C_{V_1}}{\frac{C_{V_2}P_{V_2}}{P_{V_2}+PV_3} + \frac{C_{V_3}P_{V_3}}{P_{V_2}+PV_3}}$$

$$= q\Big(\frac{P_{V_1}}{1-P_{V_1}}\Big)\frac{C_{V_1}}{C_{V_2}\Big(\frac{P_{V_2}}{P_{V_2}+PV_3}\Big) + C_{V_3}\Big(\frac{P_{V_3}}{P_{V_2}+PV_3}\Big)}$$

At this point, we recognize that $\frac{P_{V_2}}{P_{V_2}+P_{V_3}}$ is equivalent to $P_{V_2}$ if variants 2 and 3 are the only variants in that system. Assuming a 2-variant system with variants 2 and 3 only, we can use the results of the previous section to write $P_{V_2}$ as a function of the observed odds in this system:

$$P_{V_2} = \frac{q_{V_{2,23}}\text{odds}^*_{V_{2,23}}}{1 + q_{V_{2,23}}\text{odds}^*_{V_{2,23}}}$$

where $\text{odds}^*_{V_{2,23}}$ is the observed odds of $V_2$ in this 2-variant system with $V_2$ and $V_3$ (i.e., $\frac{P^*_{V_2}}{P^*_{V_3}}$) and $q_{V_{2,23}}$ is the correction factor in this system (which we know to be equal to $\frac{C_{V_3}}{C_{V_2}}$). Therefore, we can continue our calculation of $q_{V_{1,123}}$ by substituting these values as follows:

$$\frac{P_{V_1}}{1-P_{V_1}} = q\Big(\frac{P_{V_1}}{1-P_{V_1}}\Big)\frac{C_{V_1}}{C_{V_2}\frac{q_{V_{2,23}}\text{odds}^*_{V_{2,23}}}{1+q_{V_{2,23}}\text{odds}^*_{V_{2,23}}} + C_{V_3}\frac{q_{V_{3,23}}\text{odds}^*_{V_{3,23}}}{1+q_{V_{3,23}}\text{odds}^*_{V_{3,23}}}}$$

$$q = \frac{1}{C_{V_1}}\Big(C_{V_2}\frac{q_{V_{2,23}}\text{odds}^*_{V_{2,23}}}{1 + q_{V_{2,23}}\text{odds}^*_{V_{2,23}}} + C_{V_3}\frac{q_{V_{3,23}}\text{odds}^*_{V_{3,23}}}{1 + q_{V_{3,23}}\text{odds}^*_{V_{3,23}}}\Big)$$

#### 1.3 Correction factor in an n-variant system

We can extend the conclusion above to derive a formula for the correction factor $q$ in a system with $n$ variants:

$$q_{V_{1,12..n}} = \frac{1}{C_{V_1}}\Big(C_n\frac{q_{V_{n,2..n}}(odds^*_{V_{n,2..n}})}{1 + q_{V_{n,2..n}}(odds^*_{V_{n,2..n}})} + C_{n-1}\frac{q_{V_{n-1,2..n}}(odds^*_{V_{n-1,2..n}})}{1 + q_{V_{n-1,2..n}}(odds^*_{V_{n-1,2..n}})} + ... + C_2\frac{q_{V_{2,2..n}}(odds^*_{V_{2,2..n}})}{1 + q_{V_{2,2..n}}(odds^*_{V_{2,2..n}})}\Big)$$

The exact value of $q$, and therefore the true value of $P_{V_1}$, can be calculated recursively given only the coefficients of detection and observed proportions of the variants in the population.

### 2 Sample size calculations with continuous surveillance

#### 2.1 Sample size calculation for varaint detection on or before time $t$

The probability of detecting a variant (i.e., generating one or more high quality sequences indicating a patient was infected by this variant) on or before time $t$ is equal to one minus the probability of not detecting it at any time between $t_0$ and $t$. In other words, regardless of the time unit used, this probability can be written as:

$$\Pr(d \leq t) = 1 - \prod_{x=0}^{t} \Big[1 - \Pr(\text{detection at time } x)\Big]$$

Assuming a binomial sampling process, the probability of detection at time $x$ is equal to one minus the probability of not detecting the variant:

$$\Pr(\text{detection at time } x) = 1 - (1 - P_x)^n$$

Where $n$ is the sample size and $P_x$ is the prevalence of the variant at this time step. Therefore, we can write the probability of detecting the variant on or before time $t$ as:

$$\Pr(d \leq t) = 1 - \prod_{x=0}^{x=t} \Big[1 - (1 - (1 - P_x)^n)\Big] = 1 - \prod_{x=0}^{x=t} \Big(1 - P_x\Big)^n$$

Where $n$ is the per-time step sample size and $P_x$ is the prevalence of the variant at time $x$. This assumes the same number of samples are selected at every time step, and that the prevalence of the variant at each time step is known. We can rewrite this equation to solve for the per-time step sample size:

$$n = \frac{\ln[1 - \Pr(d \leq t)]}{\ln[\prod_{x=0}^{t}(1 - P_x)]}$$

We can approximate the value of the product with a continuous function using the Volterra product integral. For a scalar function $f$ and real values of $a$ and $b$:

$$\prod_{a}^{b} \Big(1 + f(x)dx\Big) = \lim_{\Delta x \to 0} \prod \Big(1 + f(x_i)\Delta x\Big) = \exp\Big(\int_a^b f(x)dx\Big)$$

Let $f(x) = -P_x dx$. This allows us to write the product of $1 - P_x$ as:

$$\prod_{x=0}^{t} \Big(1 + (-P_x dx)\Big) = \exp\Big(\int_0^t -P_x dx\Big) = \frac{1}{\exp(\int_0^t P_x dx)}$$

If we plug this into our per-time step sample size calculation we get:

$$n = \frac{\ln[1 - \Pr(d \leq t)]}{\ln[\frac{1}{\exp(\int_0^t P_x dx)}]} = \frac{\ln[1 - \Pr(d \leq t)]}{\ln(1) - \ln[\exp(\int_0^t P_x dx)]} = -\frac{\ln[1 - \Pr(d \leq t)]}{\int_0^t P_x dx} = -\frac{\ln[1 - \Pr(d \leq t)]}{G(t) - G(0)}$$

Where $G(t)$ is the integral of $g(t)$. In other words, $G(t)$ is the cumulative density function of the growth function ($g(t) = P_x dx$) used to model the change in the variant frequency over time.

#### 2.2 Logistic growth of variant frequency

Growth in prevalence of variants of interest (i.e., variants with some fitness advantage) are often modeled by logistic growth functions. In other words:

$$g(t) = \frac{1}{1 + ae^{-rt}}$$

Where $r$ is the per-time step growth rate and $a = \frac{1}{t_0} - 1$. The cumulative density of this probability distribution can be computed as follows:

$$\begin{aligned} G(t) &= \int \frac{1}{1+ae^{-rt}} dt = \int \Big(\frac{e^{rt}}{e^{rt}}\Big) \frac{1}{1+ae^{-rt}} dt = \int \frac{e^{rt}}{e^{rt}+a} dt \\ &= \int \frac{1}{u} \frac{du}{r} = \frac{1}{r} \int \frac{1}{u} du = \frac{1}{r} \ln |u| \\ &= \frac{1}{r} \ln |a + e^{rt}| + C \end{aligned}$$

Where $u = e^{rt} + a$.

#### 2.3 Logistic growth of variant frequency with biased detection

In a two-variant system, the observed prevalence of a particular variant of interest in a sample of high quality detected infections ($H$) is a function of the true prevalence and the ratio between coefficients of detection:

$$\text{observed frequency} = \frac{P_{V_1}}{P_{V_1} + \frac{C_{V_2}}{C_{V_1}} P_{V_2}}$$

Assuming $P_{V_1}$ can be computed at any given time step using a logistic model, we can calculate the observed frequency distribution as follows:

$$\text{observed frequency} = \frac{\frac{1}{1+ae^{-rt}}}{\frac{1}{1+ae^{-rt}} + \frac{C_{V_2}}{C_{V_1}}(1 - \frac{1}{1+ae^{-rt}})} = \frac{1}{1 + \frac{C_{V_2}}{C_{V_1}} ae^{-rt}} = \frac{1}{1 + be^{-rt}}$$

Where $b = a\frac{C_{V_2}}{C_{V_1}} = (\frac{C_{V_2}}{C_{V_1}})(\frac{1}{t_0} - 1)$.

Because in this case the observed frequency function takes the same form as the actual frequency function, we can easily calculate $G^*(t)$, the cumulative density of the observed variant frequency:

$$G^*(t) = \frac{1}{r} \ln |b + e^{rt}| + C$$

#### 2.4 Sample size calculation for determining variant prevalence from $t$ measurements

We can calculate the mean of multiple prevalence estimates to refine our estimate of variant frequency in a population. Using a weighted mean allows us to give more value to recent measurements. Given a particular weighting scheme, we can calculate the number of sequences needed per measurement ($n$) using the effective sample size.

Binomial theory tells use that the effective sample size of independent observations (i.e., prevalence estimates drawn from $n$ sequences) is a value such that:

$$\mathrm{Var}(\hat{\mu}) = \frac{\sigma^2}{n_{\text{eff}}}$$

Where $\mu^2$ is the mean of the estimates across all samples and $\sigma^2$ is the variance of the underlying distribution (i.e., a Bernoulli distribution, since each sequence can be either the variant of interest or not the variant of interest, with probability $p$ — the true prevalence of the variant in the population).

We also know that the the variance of each prevalence estimate is:

$$\mathrm{Var}(s_i) = \frac{pq}{n}$$

Where $n$ is the number of sequences used in that prevalence estimate, $p$ is the true prevalence of the variant in the population, and $q = 1 - p$.

If we assume that each prevalence estimate is made from the same number of sequences ($n$) and that the true prevalence is roughly equal across time points, we can calculate the variance of the mean of multiple prevalence estimates given a particular weighting scheme:

$$\mathrm{Var}(\hat{\mu}) = \mathrm{Var}\,\big(\sum_{i=1}^{t} w_i s_i\big) = \mathrm{Var}(s_i)\sum_{i=1}^{t} w_i^2 = \frac{pq}{n}\sum_{i=1}^{t} w_i^2$$

Where $w$ is the weight of the prevalence estimate from timepoint $i$ and $t$ is the total number of timepoints considered in each weighted estimate.

If the weights do not sum to one, we need to scale this estimate by the sum of the weights:

$$\mathrm{Var}(\hat{\mu}) = \mathrm{Var}\,\big(\frac{\sum_{i=1}^{t} w_i s_i}{\sum_{i=1}^{t} w_i}\big) = \frac{1}{\big(\sum_{i=1}^{t} w_i\big)^2}\,\mathrm{Var}\,\big(\sum_{i=1}^{t} w_i s_i\big) = \frac{\sum_{i=1}^{t} w_i^2}{\big(\sum_{i=1}^{t} w_i\big)^2}\big(\frac{pq}{n}\big)$$

Since we know that $\sigma^2 = pq$:

$$n_{\text{eff}} = n\frac{\big(\sum_{i=1}^{t} w_i\big)^2}{\sum_{i=1}^{t} w_i^2}$$

### Supplemental Text 2: Worked Example

In the text below, we apply our sampling methodology to prepare for the emergence of a new variant of a SARS-CoV-2-like pathogen into a fictional population, based on current whole genome sequencing capacity and experience with previous variants of concern. All calculations described can be easily performed using our Excel spreadsheet (Supplemental Data 1) or R package, `phylosamp`.

Following the diagram shown in Figure 1, the first step in applying our method is to determine the population of interest. In this example, we'll assume we are interested in tracking variants of Pathogen X in a small country with a well-defined population. Next, we need to identify the key question we are trying to answer with our surveillance scheme. In this example, we will assume we are interested in calculating the sample size needed to detect the emergence of a new variant of Pathogen X by the time it reaches a frequency of 1% across all infected individuals in our country. In other words, we will focus on **variant detection**. Finally, we need to identify the sampling frequency we have the capacity to maintain. In our case, we'll assume we want to develop a weekly sampling scheme, in which pathogen samples collected over a 7-day period are sequenced in weekly batches (i.e., **periodic surveillance**).

Now that we've identified our surveillance goals, we need to estimate some basic parameters for our population of interest, such as the pathogen testing rate, the sensitivity of the tests used, etc. However, since these values may vary by pathogen variant, we need to explore and estimate these parameters in a variant-specific context. In the current implementation of the sample size calculation methodology described herein, the specific parameters we will need to consider are (see Table 1): the variant-specific asymptomatic rate, the asymptomatic and symptomatic testing rates, the variant-specific testing sensitivity using currently available technologies, the variant-specific sampling success rate (i.e., the expected number of samples of high enough quality for variant characterization by whole genome sequencing), and the sequencing success rate. Let's consider each of these parameters in turn.

#### 1 Variant-specific parameter estimation

**The asymptomatic rate** ($\psi$). So far, epidemiologists have determined that the asymptomatic rate of Pathogen X has ranged from 30-40%. The currently circulating variant has an asymptomatic rate of 30%. Given this, we want to plan for variants that could have asymptomatic rates ranging from 25-45%. In Figure S2, we can see that a lower asymptomatic rate causes enrichment of the variant in sampled infections; conversely, a higher asymptomatic rate would artificially deplete samples belonging to the variant of interest in the pool of detected infections. Therefore, an asymptomatic rate of 25% represents the least conservative scenario (since enrichment of a variant of interest would mean fewer sequences are required to detect it) while an asymptomatic rate of 45% represents the most conservative scenario.

**The testing rate** ($\tau$). Given the widespread availability of rapid antigen tests for Pathogen X, we assume that only 50% of symptomatic infections (of any variant) are tested, and only 10% of asymptomatic infections are detected and samples sent to national public health laboratories. We anticipate that testing rates could drop as low as 40% (symptomatic) / 5% (asymptomatic) as the population becomes increasingly desensitized to disease spread. Because of the complex relationship between testing rates and sampling bias, we'll explore these two scenarios independently when performing sample size calculations.

**The testing sensitivity** ($\phi$). The current gold-standard PCR test for Pathogen X has a sensitivity of 95% for the currently circulating variant. Historical data shows this rate has changed very little between variants. However, to account for the possibility that a future variant may significantly change the viral load present in patient samples or mutate in such a way that tests temporarily become less effective (until an updated PCR target can be developed), we will perform sample size calculations assuming no change in sensitivity (least conservative scenario) as well as a drop in sensitivity down to 90% (most conservative scenario, see Figure S2).

**The sampling success rate** ($\gamma$). In many laboratory settings, viral load is measured for each sample by qPCR prior to sequencing, and the results are used to select samples for sequencing. Only sequencing the highest quality samples ensures the sequencing process is maximally cost effective. For the sake of example, we will assume that the sampling success rate is expected to be the same across all potential variants. However, we can imagine that a variant with a lower sampling success rate would require additional sampling for accurate detection.

**The sequencing success rate** ($\omega$). Not all samples selected for sequencing will produce high quality genomes that can be used for variant characterization. We assume that sequencing success is fixed across all variants, as the factors that affect sequencing success are not independent of those affecting infection detection and sample quality. In the national laboratory of our country of interest, the sequencing success rate is 80%.

#### 2 The coefficient of detection ratio

Once we have estimated the parameter ranges of interest, we can calculate the *coefficient of detection* in the most and least conservative scenarios. We can do this using the Excel spreadsheet provided as Supplemental Data 1 or using the `vartrack_cod_ratio()` function of the R package `phylosamp`.

When calculating the coefficient of detection, keep in mind that the $\gamma$ parameter can be left out (R package) or set to 1 (under "Average proportion of samples below Ct threshold") for both the variant of interest and general population parameters, since (as discussed above) we are assuming that this parameter does not change between variants. In the least conservative scenario as described above, the testing sensitivity $\phi$ also does not differ between potential new variants and the currently circulating pathogen population.

We can provide the remaining parameters as follows. (Note that $V_1$ represents the future variant we want to capture and $V_2$ parameters correspond to the general pathogen population.)

- Least conservative scenario assuming higher testing rate: $\psi_{V_1} = 0.25$, $\psi_{V_2} = 0.3$, $\tau_a = 0.1$, $\tau_s = 0.5 \implies$ coefficient of detection ratio $= 1.053$.
- Least conservative scenario assuming lower testing rate: $\psi_{V_1} = 0.25$, $\psi_{V_2} = 0.3$, $\tau_a = 0.05$, $\tau_s = 0.4 \implies$ coefficient of detection ratio $= 1.059$.
- Most conservative scenario assuming higher testing rate: $\psi_{V_1} = 0.45$, $\psi_{V_2} = 0.3$, $\phi_{V_1} = 0.90$, $\phi_{V_2} = 0.95$, $\tau_a = 0.1$, $\tau_s = 0.5 \implies$ coefficient of detection ratio $= 0.798$.
- Most conservative scenario assuming lower testing rate: $\psi_{V_1} = 0.45$, $\psi_{V_2} = 0.3$, $\phi_{V_1} = 0.90$, $\phi_{V_2} = 0.95$, $\tau_a = 0.05$, $\tau_s = 0.4 \implies$ coefficient of detection ratio $= 0.779$.

Given these results, we can move forward to sample size calculations with two values of the coefficient of detection ratio to test: 0.779 (most conservative scenario) and 1.059 (least conservative scenario).

#### 3 Sample size calculations

Once we have determined the range of scenarios we'd like to explore, we can perform sample size calculations using the appropriate tab/function of the Excel spreadsheet or R package, respectively. As our aim is to ensure variant detection using a periodic sampling strategy, we need to use the “Detect (Periodic) - Sample Size” tab of the Excel spreadsheet or the `vartrack_samplesize_detect()` function of the `phylosamp` R package.

In both cases, there are a few more parameters we need to provide:

**The desired probability of detection** ($prob$). We can again select a parameter range to explore for our sample size calculations. In our case, we want to ensure a good chance of detecting a new variant of Pathogen X when it enters our country, so we will explore probabilities of detection between 75% (least conservative) to 95% (most conservative).

**The desired variant prevalence** ($p_{V_1}$). As stated above, we want to ensure we catch any variant by the time it has reached 1% prevalence in the population of infected individuals.

**Initial variant prevalence** ($p_0$). The method we will use for sample size calculations assumes logistic growth of any new variants of concern, with a starting prevalence and growth rate that can be specified. This initial prevalence depends on the number of simultaneous variant introductions into our country of interest as well as the total infected population size. Over the last year, we have observed a total of between 5,000 and 10,000 total cases of Pathogen X in our country at any given time, and we expect the number of cases to be similar if a new variant is introduced. Because of the complex relationship between initial prevalence and the shape of the logistic growth curve, we will estimate the required sample size in two scenarios: (1) if a new variant could be introduced via a single index case, at a time when nearly 10,000 people are infected (initial prevalence $= 1/10000$); and (2) if 5 different travelers are be infected by a new variant and bring it into the country in the same week, at a time when only 5,000 individuals are infected (initial prevalence $= 5/5000 = 1/1000$).

**Logistic growth rate** ($r$). We also need to estimate a variant growth rate over time. Based on historical data of Pathogen X, we know that a recently introduced variant may grow as slowly as 0.1x/day (least conservative, as fewer samples are needed to ensure we catch the variant before it goes above 1% frequency) or as quickly as 0.2x/day (most conservative).

We now have all of the values we need to estimate the sample size needed for detecting a variant by the time it reaches 1% in the population, assuming weekly periodic sampling. Additionally, it is important to remember that the number of required **sequences** is not the same as the number of required **samples**, because of the sequencing success rate ($\omega$) discussed above. Both the Excel spreadsheet and `phylosamp` R package output the number of samples required, taking into account that not all samples selected for sequencing will result in high quality samples suitable for variant characterization:

- Least conservative scenario with low initial prevalence: $prob = 0.75$, $p_{V_1} = 0.01$, $p_{0_{V_1}} = 1/10000$, $r_{V_1} = 0.1$, $\omega = 0.8$, coefficient of detection $= 1.059 \implies$ 112 samples should be sequenced per week (16 per day)
- Least conservative scenario with high initial prevalence: $prob = 0.75$, $p_{V_1} = 0.01$, $p_{0_{V_1}} = 1/1000$, $r_{V_1} = 0.1$, $\omega = 0.8$, coefficient of detection $= 1.059 \implies$ 119 samples should be sequenced per week (17 per day)
- Most conservative scenario with low initial prevalence: $prob = 0.95$, $p_{V_1} = 0.01$, $p_{0_{V_1}} = 1/10000$, $r_{V_1} = 0.2$, $\omega = 0.8$, coefficient of detection $= 0.779 \implies$ 567 samples should be sequenced per week (81 per day)
- Most conservative scenario with high initial prevalence: $prob = 0.95$, $p_{V_1} = 0.01$, $p_{0_{V_1}} = 1/1000$, $r_{V_1} = 0.2$, $\omega = 0.8$, coefficient of detection $= 0.779 \implies$ 679 samples should be sequenced per week (97 per day)

Based on these calculations, we need to be sequencing between 112 and 679 samples per week in order to detect a new variant by the time it reaches 1% in the population. As this is a rather wide range, we can use the reverse functionality of the sample size calculation method to determine the probability of detecting a variant given a fixed number of samples and most conservative parameter values.

### 4 Estimating the probability of detection

Given the recommendation of 112-679 samples per week, the government of our country of interest has decided that funding will be allocated to support sequencing of 200 Pathogen X samples per week. Given our most conservative scenario of a coefficient of detection of 0.779 and a growth rate of 0.2, we can use the "Detect (Periodic) - Confidence" tab of the Excel spreadsheet (or the `vartrack_prob_detect()` function in the `phylosamp` package) to calculate the probability of detecting a variant before it crosses the 1% prevalence threshold in the population.

In both the high and low initial prevalence scenarios, the probability of detection (assuming roughly 28 samples selected per day, to be sequenced in weekly batches) remains above 57% even using the most conservative parameters. Furthermore, the probability of detecting a new variant by the time it reaches 2% in the population is approximately 85% in both scenarios, with numbers approaching 99% chance of detection before the variant hits 5% prevalence. These values may be sufficient for country officials to feel confident in their ability to detect a variant soon after it is introduced regardless of its biological properties; if it is not, the calculations can simply be repeated with a higher number of weekly samples.

Of course, there are many assumptions that underlie these calculations, the most obvious being that the weekly batch of samples for sequencing are assumed to be well-distributed across the days of the week, and that they capture all regions or ports of entry into the country. Even so, this method provides sampling guideposts that can be applied in a variety of settings. For example, it is clear from the simple calculations above that 100 samples per week would be unlikely to be particularly informative for detecting new variants early and with high confidence.

Although the example provided here focuses on the question of detection with periodic sampling, the same principles (though different functions/spreadsheet tabs) can be applied to a cross-sectional sampling scheme and/or estimating variant prevalence. The section on the coefficient of detection remains identical, and only the sampling calculations need to be updated to suit the surveillance goals.
